# Comparative analysis of post-vaccination anti-spike IgG antibodies in old Nursing Home Residents and in middle-aged Healthcare workers

**DOI:** 10.1101/2021.08.03.21261014

**Authors:** H. Jeulin, D. Craus, C. Labat, A. Benetos, on behalf of the “Lorraine Nursing Homes network on COVID-19 vaccine response”

**Affiliations:** Université de Lorraine, CNRS, LCPME, F-54000 Nancy, France; Laboratoire de Virologie, CHRU de Nancy Brabois, F-54500 Vandœuvre-lès-Nancy, France; Maison Médicale F-54110 Rosières-aux-Salines, France; Université de Lorraine, Inserm, DCAC, F-54500 Vandœuvre-lès-Nancy, France; Université de Lorraine, CHRU-Nancy Brabois, Department of Clinical Geriatrics F-54500 Vandœuvre-lès-Nancy France

**Keywords:** nursing homes, very old, vaccination, COVID-19

## Abstract

The current consensus is that 2 doses of mRNA vaccines against SARS-CoV-2 are needed for people without COVID-19 history, while for those who had suffered COVID-19, a single dose may be enough to achieve high levels of immunization. This consensus has been based on results obtained in middle-aged populations, whereas only few data exist for the oldest and most frail adults such as nursing home residents (NH-Res) that is, the population most vulnerable to develop severe forms of COVID-19.

In this study, we studied the anti-SARS-CoV-2 IgG(S) level of NH-Res and healthcare workers (HCWs) with or without a history of COVID-19 infection taking into account the time since immunization (COVID-19 and/or vaccination). 654 subjects were analyzed: 397 NH-Res (median age 88, IQR 82-93 years, 75% women) and 257 Health Care Workers (HCWs, median 46, IQR 38-54 years, 81% women). NH-Res and HCWs were classified in one of the following 3 groups: No-COVID history and 2 vaccine shots (COV-NO/2VACC); Yes-COVID history and 1 (COV-YES/1VACC) or 2 (COV-YES/2VACC) vaccine injections.

The time-related decrease in IgG (S) in subjects without COVID-19 history, SARS-COV-2 serology would be negative in HCWs approximately 220 days and in residents 180 days after vaccination. This time-related decrease was much slower in those with history of COVID. NH-Res belonging into the COV-NO/2VACC and the COV-YES/1VACC groups showed lower IgG (S) levels than the same groups of HCWs (for both groups, p<0.0001), whereas in the group COV-YES/2VACC, IgG (S) levels were similar in NH-Res and HCWs (p=0.88). These results remained unchanged after adjustment for age and duration since immunization. Thus, in NH-Res, 2 vaccine shots were associated with a more pronounced immune response, whereas in HCWs, 1 or 2 vaccine shots in patients with COVID-19 history did make any difference. These results indicate significant differences in mRNA vaccination between NH-Res and middle-aged controls, and could contribute to the specification of vaccine policy in this very old, frail population.

## Introduction

The development of vaccines against SARS-CoV-2, allows the possibility for massive immunization of the population and the control of coronavirus disease 2019 (COVID-19). mRNA vaccines have been used in hundreds of millions of people showing high efficacy and safety profiles (1,2) even in older patients (3). Several public health and political authorities have raised the question of the number of doses of vaccines required to achieve high levels of immunization. The current consensus is that 2 doses are needed for people without a history of COVID-19, while for those who have suffered COVID-19, a single dose may be enough to reach the level reached by people not infected who received 2 injections (4-6). These studies were mainly carried out in young and middle-aged populations; there are however few data for the oldest and most frail adults such as residents of nursing homes (NHs) that is, the population most vulnerable to develop severe forms of COVID-19. One recent study by Blain et coll has shown that in NH residents with history of COVID-19, one injection of mRNA vaccine was followed by a significant development of anti-spike IgG (7). However, no study has assessed the magnitude of the antibody response to mRNA vaccination in very old subjects in comparison to young-middle aged individuals i.e. subjects who showed a very good response to vaccination against SARS-CoV-2.

To this end, we compared the post-vaccination anti-SARS-CoV-2 IgG(S) level in NH Residents (NH-Res) and in healthcare workers (HCWs) with or without a history of COVID-19 infection taking into account the time between the “last immune stimulation” (COVID-19 and/or vaccination) and the moment of the IgG detection.

## Methods

NH-Res of 10 NHs participating in this study (including the NH of the Geriatric Dpt of the University hospital of Nancy) and HCWs from the same NHs and the Geriatric Dpt of the University hospital of Nancy were included (see list of the participant NHs in the front page). The current analysis was coordinated by the Geriatric Dpt of the University hospital of Nancy (CHRU de Nancy). The directors and medical coordinators of these centers gave their agreement for the participation to this study. Residents and staff have been informed orally or with posters in the respective health institutions, of the possibility of using their serology results for clinical research purposes. The Ethics Committee of the Nancy CHRU hospital made the statement that this research has been carried out in accordance to current French and European ethical standards as well as The Code of Ethics of the World Medical Association (Comité d’Ethique CHRU de Nancy, decision n° 326, August 3, 2021).

This study has declared in ClinicaTrials (ClinicalTrials.gov Identifier: NCT04964024). Inclusion criteria for NH-Res and HCWs were:

- A SARS-COV-2 (anti-spike IgG) serology performed between April 1 - July 2, 2021.
- Two injections of mRNA vaccines (3-6 weeks apart) with the last injection at least 7 days before antibody detection independently of history of COVID-19. The subjects who were contaminated were identified either by a positive rt-PCR test at the time of contamination or by an anti-nucleoprotein (N) IgG detection test.
- One dose of mRNA vaccine, for people who declared history of COVID-19 and having a positive rt-PCR and/or detection of anti-nucleoprotein (N) IgG
- No objection to the use of their results for this analysis.

Anti-SARS-COV-2 IgG (S) were detected in 217 HCWs, and 132 NH-Res using the LIAISON® SARS-CoV-2 TrimericS IgG assay (Diasorin, France) (University hospital of Nancy); and in 40 HCWs and 265 NH-Res using the Anti-SARS-CoV-2 CLIA-YHLO kit (Shenzhen Yhlo Biotech Co., Ltd.), (Atoutbio Laboratories Nancy;). Preliminary tests (External Quality Assessment) did not show significant differences in the results obtained with these 2 methods. Also, the IgG (S) values obtained in the present study did not show any difference between the two methods after adjusting for confounders. For both methods the range was: 1->800 AU/mL. Positivity threshold was 10 and 13 AU/ml for Yhlo and Liaison XL methods, respectively.

In each of the 2 cohorts (NH-Res and HCWs), the following 3 groups were identified according to the history of COVID-19 and the vaccination status:

- No-COVID and 2 vaccine injections (COV-NO/2VACC)
- Yes-COVID and 1 vaccine injection (COV-YES/1VACC)
- Yes-COVID and 2 vaccine injections (COV-YES/2VACC)

Descriptive results are presented as medians (IQR) or percentage. Anti-SARS-CoV-2 IgG (S) levels were compared among the 3 groups in each population and between residents and health professionals in each of the 3 groups. Due to the absence of normal distribution of the IgG (S) values, comparisons between the different groups were performed after logarithmic transformation. Comparisons were performed with ANOVA tests. Age and time (Δ) between the last immune stimulation (COVID-19 or vaccine) and the antibody detection were used in the adjusted models. When the ANOVA test was statistically significant, a post-hoc Tukey-Kramer’s test was used to test the 2 by 2 differences. Multiple regression analyses were also conducted in order to test the role of the number of vaccine injections (1 vs.2) in residents and health professionals with history of COVID-19. Pearson correlation was used to study the association between IgG (S) values and time since immunization in COV-NO/2VACC in NH-Res and in HCWs. A P <0.05 was considered significant.

## Results

A total of 654 subjects were included: 397 NH-Res (median age 88, IQR 82-93 years, 75% women) and 257 HCWs (median 46, IQR 38-54 years, 81% women). Table 1 shows the main characteristics of the Covid-19/Vaccine groups in each one of these 2 populations.

**Table 1:**
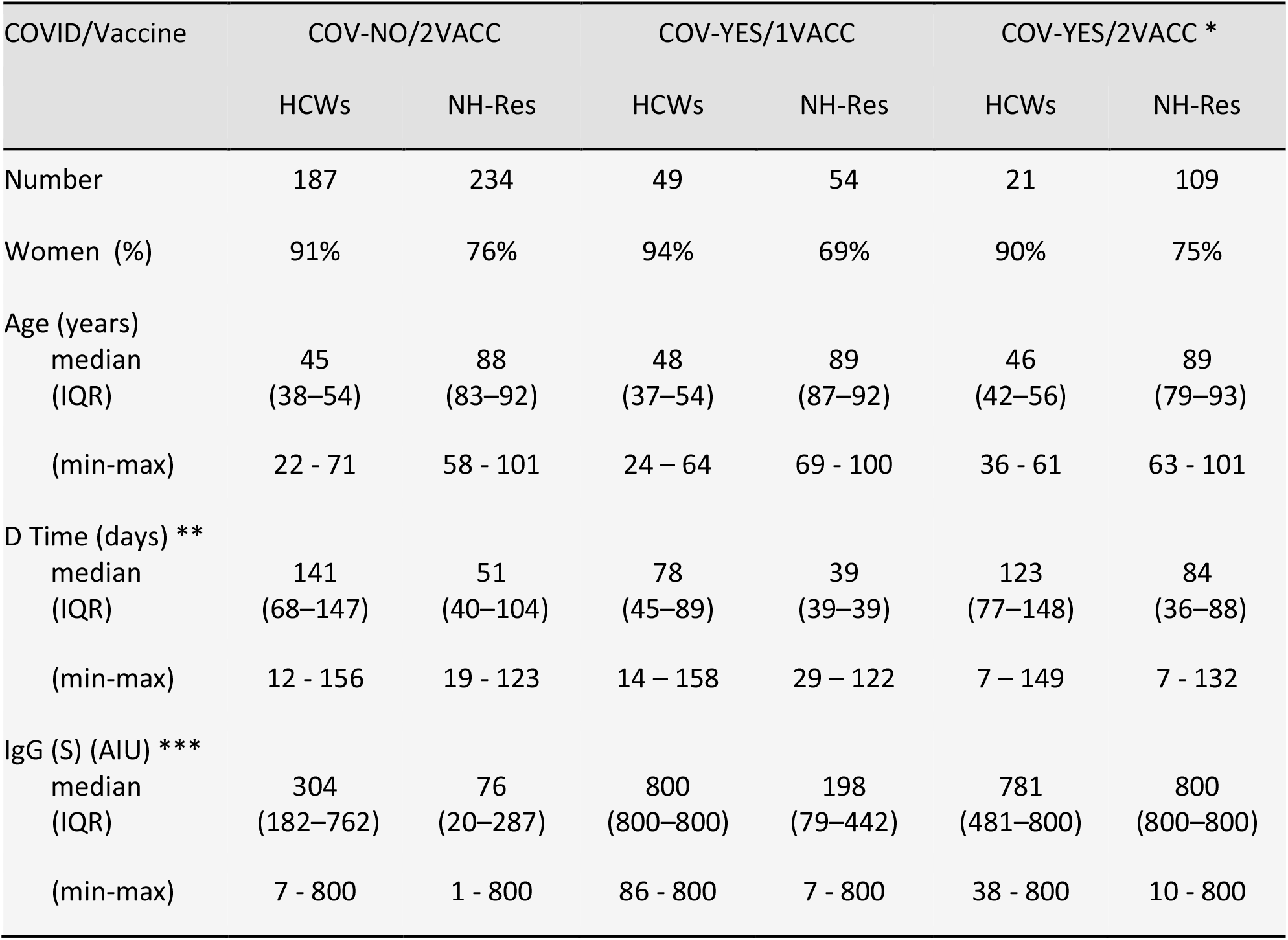
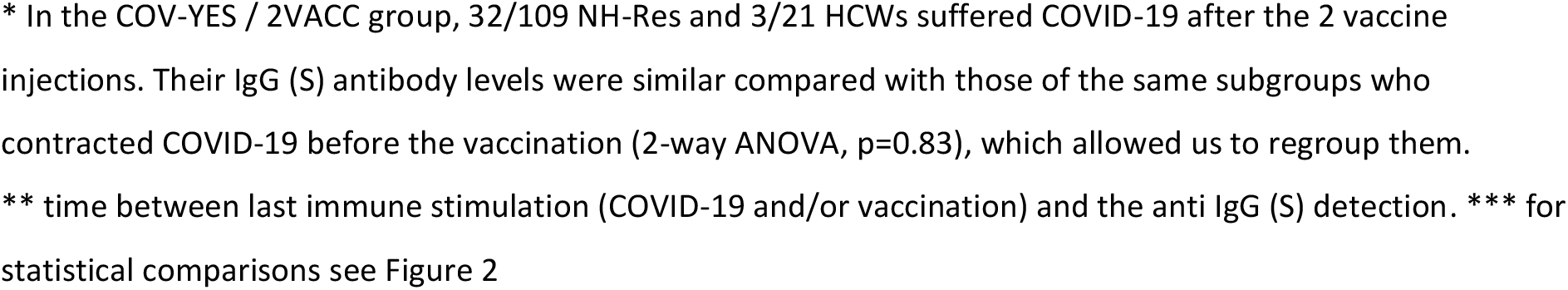
Sex distribution, age (media, IQR), time from the last immune stimulation and IgG (S) levels in the 3-COVID-19/vaccine groups of HCWs and NH-Res.

| COVID/Vaccine | COV-NO/2VACC |  | COV-YES/1VACC |  | COV-YES/2VACC * |  |
| --- | --- | --- | --- | --- | --- | --- |
|  | HCWs | NH-Res | HCWs | NH-Res | HCWs | NH-Res |
| Number | 187 | 234 | 49 | 54 | 21 | 109 |
| Women (%) | 91% | 76% | 94% | 69% | 90% | 75% |
| Age (years) |  |  |  |  |  |  |
| median | 45 | 88 | 48 | 89 | 46 | 89 |
| (IQR) | (38–54) | (83–92) | (37–54) | (87–92) | (42–56) | (79–93) |
| (min-max) | 22 - 71 | 58 - 101 | 24 – 64 | 69 - 100 | 36 - 61 | 63 - 101 |
| D Time (days) ** |  |  |  |  |  |  |
| median | 141 | 51 | 78 | 39 | 123 | 84 |
| (IQR) | (68–147) | (40–104) | (45–89) | (39–39) | (77–148) | (36–88) |
| (min-max) | 12 - 156 | 19 - 123 | 14 – 158 | 29 – 122 | 7 – 149 | 7 - 132 |
| IgG (S) (AIU) *** |  |  |  |  |  |  |
| median | 304 | 76 | 800 | 198 | 781 | 800 |
| (IQR) | (182–762) | (20–287) | (800–800) | (79–442) | (481–800) | (800–800) |
| (min-max) | 7 - 800 | 1 - 800 | 86 - 800 | 7 - 800 | 38 - 800 | 10 - 800 |

\* In the COV-YES / 2VACC group, 32/109 NH-Res and 3/21 HCWs suffered COVID-19 after the 2 vaccine injections. Their IgG (S) antibody levels were similar compared with those of the same subgroups who contracted COVID-19 before the vaccination (2-way ANOVA, $p=0.83$ ), which allowed us to regroup them.
\*\* time between last immune stimulation (COVID-19 and/or vaccination) and the anti IgG (S) detection. \*\*\* for statistical comparisons see Figure 2

### Decrease in IgG(S) according to the time from immunization

In non-COVID subjects (both HCWs and NH-Res) time from the 2^nd^ vaccine injection was negatively associated with the IgG (S) levels (Figure 1). More precisely, in COV-NO/2VACC health professionals (n=187) this relationship was expressed with the equation: IgG (S) (AU) = 808 - 3.64 X days (R=-0.63, p<0.00001) and in residents (n=234) as follows: IgG (S) (AU) = 341 - 1.89 X days (R=-0.25, p<0.0001)

**Figure 1:**
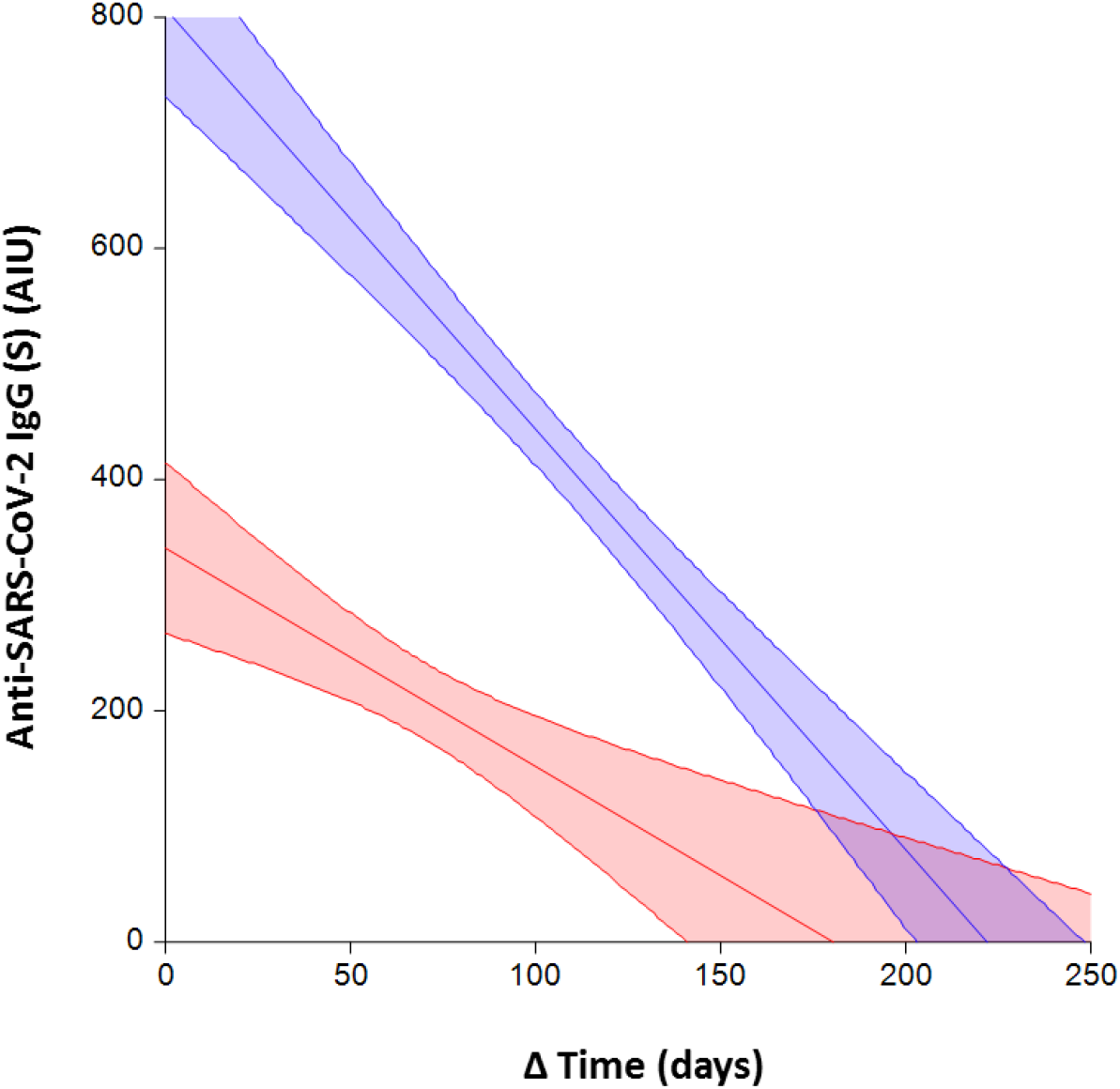
Linear correlation (CL 95%) between time (**Δ**) from the 2^nd^ injection and IgG (S) levels in HCWs (blue) and NH-Res without COVID-19 history.

In HCWs with history of COVID due to the low number of subjects in each subgroup the same analyses were conducted after grouping those with 1 (n=49) or 2 vaccine injections (n=21). This was possible since IgG responses to 1 or 2 injections were not different (see Table 1 and Figure 2). The relationship between IgG (S) and the time from last immune stimulation was expressed by the equation: IgG (S) (AU)= 853 - 1.82 X days (R= -0.35, p=0.003). In NH-Res with COVID-19 history, grouping those with 1 and 2 vaccine injections was not possible because of the different IgG (S) responses between these 2 subgroups, and therefore analyses were performed separately: In the group COV-YES/2VACC (n=109) the equation was: IgG(S) (AU) = 780 - 0.98 X days (R= -0.21, p= 0.03). In the COV-YES/1VACC (n=54) no such relationship was observed (p=0.32).

**Figure 2:**
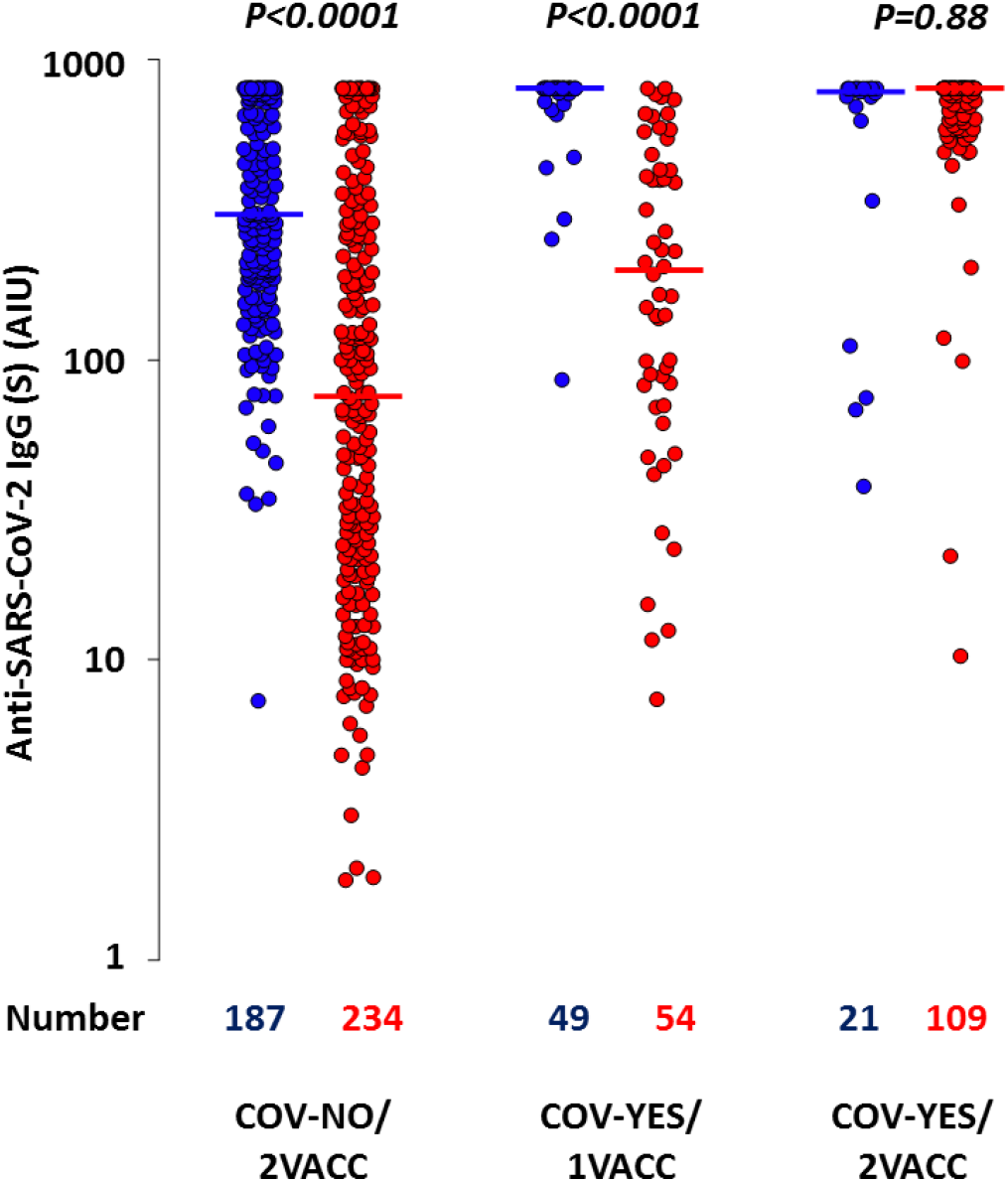
Anti-SARS-CoV-2 IgG (S) following vaccination in HCWs (blue) and NH-Res (red) in the 3 groups according to COVID-19 history and the number of vaccine doses. -COV-NO/2VACC: No history of Covid-19 and 2-vaccine injections, COV-YES/1VACC: History of Covid-19 and 1-vaccine injection, COV-YES/2VACC: History of Covid-19 and 2-vaccine injections. The horizontal lines represent median values for S-protein IgG. -p values in Figure 2 correspond to the post-hoc Tukey-Kramer’s test for comparing IgG (S) between populations of the same COVID/Vaccine status. -In HCWs the ANOVA analysis showed different IgG (S) levels among the 3 groups (p<0.0001). The 2×2 comparisons showed that IgG(S) levels were similar in the COV-YES/1VACC and COV-YES/2VACC and in both higher than in the COV-NO/2VACC group (p<0.001 and p<0.02 respectively). In the NH-Res group the ANOVA showed different IgG (S) levels among the 3 groups (p<0.0001). The 2×2 comparisons were all highly significant COV-NO/2VACC < COV-NO/1VACC < COV-NO/2VACC (all p<0.001). For both populations, adjustment for the technique used for the IgG(S) detection, age, and time from last immune stimulation did not modify the results

Age was also a significant determinant of the IgG (S) response (negative relationship) but only in subjects without history of COVID-19 (p<0.01 in HCWs and p=0.02 in NH-Res). No such relationship was observed in subjects with COVID-19 history.

### Effects of COVID-19 and number of vaccine shots on IgG(S) levels

The IgG (S) levels following vaccination are shown in Table 1 (lower panel) and in Figure 2. NH-Res of the COV-NO/2VACC and COV-YES/1VACC showed lower IgG (S) levels as compared with the same groups of HCWs (for both groups, p<0.0001). Only in the group COV-YES/2VACC IgG (S), levels in NH-Res were similar with those observed in HCWs (p=0.88) (figure 2, right).

The IgG (S) differences between NH-Res and HCWs remained unchanged after adjustment for age and time from last immune stimulation. In HCWs, IgG (S) levels were different among the 3 groups (p<0.0001); this difference concerned only the COV-NO/2VACC which showed lower IgG (S) levels compared to the COV-YES/1VACC or COV-YES/2VACC (Figure 2). In NH-Res, the 3 groups showed different IgG (S) levels (one-way ANOVA p <0.0001) and the 2×2 comparisons were all significantly different: COV-NO/2VACC < COV-NO/1VACC < COV-NO/2VACC (all p<0.001).

The role of the 2nd injection in NH-Res and HCWs with COVID history was also studied using a multiple regression analyses (including also in the model, Age and time from the last immune stimulation as independent variables). This analysis showed that the 2^nd^ vaccine injection dramatically increases IgG (S) in NH-Res by 429 ± 36 AIU (R^2^ = 43%, p<0.0001). By contrast, in HCWs, the 2^nd^ dose did not induce any modification in IgG (S) levels (p=0.23).

## Discussion

In the present study, we evaluated the SARS-CoV-2 IgG(S) antibody response to the vaccination in NHs residents i.e. in a very old (88 years at mean), frail population which is one of the most vulnerable population for having severe forms of COVID-19. Although IgG (S) levels represent just a part of the global immunity response (8), it has been shown that “compromised immune responses to the SARS-CoV-2 spike is a major trait of COVID-19 patients with critical conditions“(9). In the present study, the results obtained in the different NH-Res groups were compared with those of middle-aged HCWs with or without a history of COVID-19 infection. This comparison showed lower levels of IgG(S) in the NH-Res population in two groups: COVID-NO/VACC2 and COVID-YES/VACC1. In contrast, in the COVID-NO/VACC2, NH-Res and HCWs showed similar IgG (S) levels, indicating that the second injection of mRNA vaccine, 3 to 6 weeks after the first shot, significantly increased IgG (S) values in the very old frail subjects.

One could ask that in all these groups median IgG (S) values were clearly higher than the threshold of positivity (10-13 AU/ml) and therefore differences among the different groups are clinically not relevant. However, a recent study has shown that only sera with mean IgG (S) level > 126 AU/mL were associated with a neutralizing activity of SARS-CoV-2 on Vero E6 cells (Neutralizing titer 50 ≥ 40) (10). Therefore, it is reasonable to look for high IgG(S) levels especially in the very old, vulnerable patients.

In the present study, NH residents develop the highest antibody levels only if they have both history of COVID and 2 injections of the vaccine i.e. three SARS-CoV-2 immune stimulations, whereas in younger subjects 2 immune stimulations (COVID-NO/VACC2 or COVID-YES/VACC1) is necessary to obtain high IgG (S) levels as it has previously reported (4,5). However, even in HCWS, COVID-YES/VACC1 was associated with higher IgG(S) as compared with the COVID-NO/VACC2. Indeed, in both populations (NH-Res and HCWs), previous history of COVID-19 was associated with higher IgG (S) response to the vaccine. This is more pronounced in old age NH-Res, in which median values are particularly lower in the (COVID-NO/VACC2) as compared with all other vaccinated groups.

We also analyzed the time-related decrease in IgG (S) in the different subgroups. The obtained results allowed us to make some estimation about the time after vaccination for a serology to become negative. Following this estimation, in subjects without COVID history, IgG (S) levels would be negative in HCWs approximately 220 days and in residents 180 days after vaccination.

For subjects with history of COVID-19, a longer time delay is estimated: about 470 days, from vaccination in HCWs and for NH-Res we did not observe a significant decrease in the IgG level during the follow-up after 2 post-COVID vaccine shots. These results are in accordance with previously described data showing that very old people can elicit durable SARS-CoV-2 spike specific IgG antibodies following COVID infection (11). However, it is important to define a vaccine strategy in order to obtain sufficient neutralizing antibodies level in order to guarantee a long term sufficient immunization

One major limitation of our study is its cross-sectional character and therefore results have to be interpreted with a lot of precaution.

## Conclusions

The present results could suggest that in NH-Res residents with a history of COVID, a double vaccination provides the highest IgG (S) levels and therefore the strategy of a single vaccine shot should be reconsidered for this very old frail population. The clinical meaning of our results concerning the time-related decrease in IgG (S) need to be investigated in large longitudinal studies, analyzing the long term post vaccination neutralization capacity and precise the need of a booster vaccine shot.

## Data Availability

no

